# Increased Prevalence of Oxytocin Receptor Gene Variant in Functional Neurological Disorder: A Preliminary Case-Control Study

**DOI:** 10.1101/2024.11.26.24317955

**Authors:** Samantha Weber, Natascha Stoffel, Lucía Trinidad Rey Álvarez, Juan Ansede-Bermejo, Raquel Cruz, Álvaro Del Real Bolt, Janine Bühler, Ángel Carracedo, Selma Aybek

## Abstract

**Background:** Current models on functional neurological disorders (FND) propose a multifactorial origin. Recent studies identified potential biological vulnerability factors – such as a reduced limbic volume or an altered stress response. These findings highlight the need to investigate a potential genetic contribution to the biological vulnerability to FND.

**Method:** Eighty-five mixed FND patients and seventy-six healthy controls (HC) were genotyped for ten single nucleotide polymorphisms within seven genes associated with the stress system. For the genetic variant that was found to be associated with FND, further associations to structural brain alterations were investigated using a region-of-interest approach. Regions were previously selected based on their biological involvement and as a vulnerability for FND.

**Results:** A significant association between the diagnosis of FND and the rs53576 of the oxytocin receptor (*OXTR*) gene was found. A significant association between decreased right insular volumes and rs53576 (*OXTR*) was identified in FND patients. In female patients, the rs53576 (*OXTR*) was associated with a reduced bilateral amygdalar volume.

**Conclusion:** These preliminary results suggest a genetic contribution to the biological vulnerability for FND involving the oxytocinergic system, and (sex-specific) structural changes in insula and amygdala.

## INTRODUCTION

Functional neurological disorders (FND) comprise diverse neurological symptoms, which have been attributed to underlying functional and structural brain alterations[1] with contemporary models aiming at integrating a multifactorial origin of FND by means of a stress-diathesis model.[2] To better understand FND in a multifactorial framework, there is a need to unveil how other vulnerability factors (e.g., genetic variants) could play a role. The research on the genetic contribution towards FND is in its infancy. Preliminary results in FND showed that risk allele carriers of the tryptophan-hydroxylase 2 (*TPH2*, involved in the biosynthesis of serotonin) polymorphism rs4570625 had an earlier onset of FND and altered amygdalar functional connectivity in relation to childhood trauma.[3] We could previously partially replicate these findings, as a significant association between *TPH2* and FND symptom severity was found along with the *TPH1* genetic variant rs1800532 associated with worse clinical outcome for patients with FND.[4] Another recent study with a subpopulation of the same cohort focused on oxytocin’s involvement specifically, detecting an interaction of genotype and methylation depending on group; in the FND population, the A allele of *OXTR* SNP rs53576 was associated with higher methylation levels at *OXTR*, which was not the case for healthy controls (*Weber & Stoffel, 2024, accpted for publication*). In this short report, we present the genetic results of the same cohort of the above-mentioned study, where we analyzed a set of single-nucleotide polymorphisms (SNPs) and further investigated the association between the significantly associated SNPs with structural brain alterations in FND. The SNPs were a priori selected based on their potential relevance for the pathophysiology in FND, as were the structural regions-of-interest.

## METHODS

The study was conducted at the University Hospital Inselspital Bern, Switzerland. Eighty-six mixed FND patients and seventy-six healthy controls (HC) of Caucasian origin were recruited. All procedures were performed in compliance with relevant institutional guidelines and have been approved by the local Ethics Committee of the Canton Bern (DRKS00012992). All subjects provided written informed consent. For further details, we refer the reader to our previous work.[2,4–6] The genotyping was conducted at the Spanish National Center for Genotyping (CeGEN, Santiago de Compostela, Spain) and is detailed in the Supplementary Material.

Statistical analyses were performed using *R* software (version 4.2.1.). Alpha-level was set at *P*<0.05. We selected seven genes, from which 10 SNPs were analysed including brain-derived neurotrophic factor (*BDNF*) dopamine receptors (*DRD2*; *DRD4*), FK506 binding protein 5 (*FKBP5*), tryptophan hydroxylase (*TPH1; TPH2*) and oxytocin receptor (*OXTR*), supplementary table 1, 2. Genotype call rate, minor allele frequency (MAF), and Hardy-Weinberg equilibrium (HWE) using chi-squared test were calculated as quality control procedure.

In a first analysis, we investigated the genetic association with the diagnosis of FND in a between-group analysis. For each SNP, a logistic regression was performed using SNPassoc[7] in *R*, including age, sex, depression, trait-anxiety and experience of childhood trauma as covariates (see Supplementary Material). Three different models of inheritance were considered: co-dominant, dominant, and recessive. Odds ratios (OR) and confidence intervals (CI) were calculated to evaluate the significance of the association between group and genotype. An additional model was implemented including an interaction term between the SNPs and childhood trauma. Due to the high prevalence of FND in females[8] and the different impact of biological sex on genetics and endocrinology,[9] analyses were repeated in females only.

In a second analysis, acting on the assumption of a biological predisposition to FND, we investigated significantly associated genotypes with brain volume (analysis details in Supplementary Material). Based on our previous work on oxytocin (*Weber & Stoffel, 2024, accpted for publication*) and biological vulnerability factors,[2] we selected the hippocampus, amygdala, and the insula as regions-of-interest. For each brain region, we investigated the association between genotype and brain volume in FND patients and in HC. We included age, sex, depression, trait-anxiety, total CTQ score and total intracranial volume (TIV) as covariates of no interest.

The data are not publicly available but can be shared upon request.

## RESULTS

Genomic data from 85 FND patients and 76 healthy controls were included. Mixed symptom types were present in FND patients including motor (ICD-10[19]; F44.4) and sensory symptoms (F44.6), functional seizures (F44.5), mixed symptom type (F44.7), and persistent postural-perceptual dizziness (PPPD) (clinical details in supplementary table 3). Patients had significantly higher depression and anxiety scores (*P*<0.0001) and showed higher scores in childhood emotional and physical neglect and abuse (*P*<0.03) compared to HC, supplementary table 4.

Genotype success rate was 100% for all subjects, and SNPs were in HWE (*P*>0.05), supplementary table 2. Using a logistic regression, we identified a significant association between FND and rs53576 in the *OXTR* gene (recessive model: OR=4.17, CI=[1.31–13.23], *P*=0.012; codominant model: OR_A/A_=3.16, CI_A/A_=[0.09–11.10], OR_G/A_=0.62, CI_G/A_=[0.26–1.47], *P*=0.023), indicating that AA carriers may be more vulnerable to FND. When repeating the analyses in females only, results remained significant, Table 1. None of the other SNPs showed a significant association with FND.

**Table 1.** Association analysis between SNP and FND.

| Gene | SNP ID | Model | Genotype | Full sample |  |  |  | Females only |  |  |  |
| --- | --- | --- | --- | --- | --- | --- | --- | --- | --- | --- | --- |
|  |  |  |  | Cases |  | OR<br>[95% CI] | P-value | Cases |  | OR<br>[95% CI] | P-value |
|  |  |  |  | FND | Controls |  |  | FND | Controls |  |  |
| *OXTR* | rs53576 | Recessive | G/G-G/A | 68 | 70 | 1.00 | 0.012 | 49 | 50 | 1.00 | 0.022* |
|  |  |  | A/A | 17 | 6 | 4.17 [1.31–13.23] | * | 14 | 5 | 4.21 [1.17–15.11] |  |
|  | rs53576 | Codominant | G/G | 29 | 28 | 1.00 | 0.023 | 20 | 20 | 1.00 | 0.044* |
|  |  |  | G/A | 39 | 42 | 0.62 [0.26–1.47] | * | 29 | 30 | 0.57 [0.19–1.75] |  |
|  |  |  | A/A | 17 | 6 | 3.16 [0.90–11.10] |  | 14 | 5 | 3.09 [0.75–12.75] |  |

Abbreviations: SNP: Single-nucleotide polymorphism, *OXTR*: Oxytocin receptor Significance code: \* $P<0.05$

In FND patients only, a significant association was found between the right insular volume and the rs53576 of the *OXTR* gene (dominant model, β*=-*0.37, se=0.1, *P*=0.04) with minor A-allele carriers presented smaller brain volumes. A nominal association was identified in female FND patients, but not in the whole cohort nor HC, for the bilateral amygdala and the rs53576 of the *OXTR* gene (recessive model, left amygdala β=-0.05, se=0.04 *P*=0.02, right amygdala β=-0.04, se=0.03, *P*=0.046). This result indicated smaller brain volumes of the bilateral amygdala in female patients with FND that are homozygous for the *OXTR* rs53576 A-allele. No other significant associations were detected between the genetic component and the selected brain regions, figure 1. Results are detailed in supplementary table 5.

**Fig 1:**
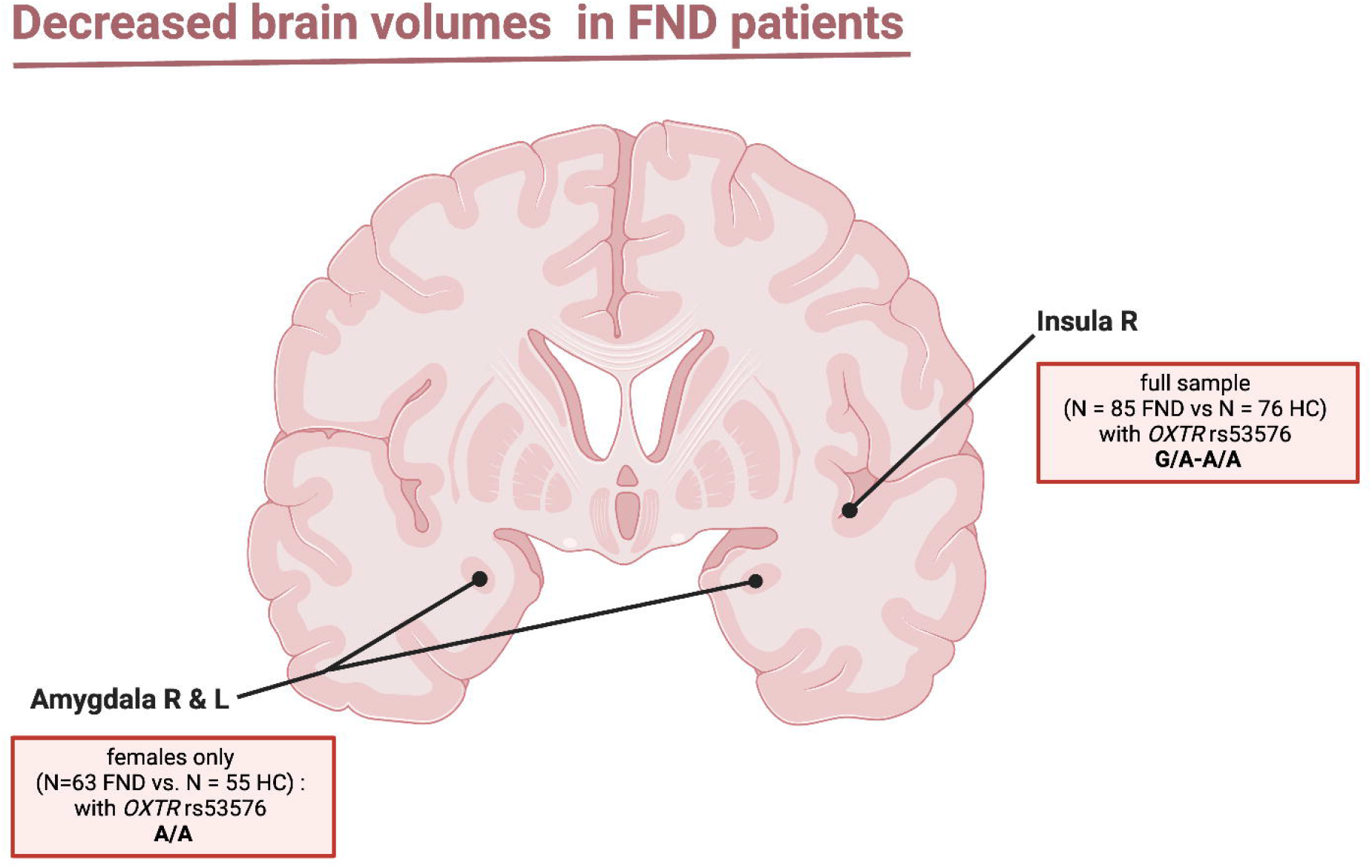
Illustrations of findings of genetic association with brain volumes in FND patients. The panel depicts genetic associations in brain regions showing decreased volumes for FND compared to healthy controls given the allelic distribution at the *OXTR* SNP rs53576. Illustration has been created in BioRender (Stoffel, N. (2025) https://biorender.com/m60f230).

## DISCUSSION

We identified a significant association between the minor allele of the rs53576 *OXTR* SNP and FND, indicating that carriers of this allele might be at higher risk for FND. This variant has been reported to be associated with stress predisposition, with A-allele carriers having a higher sensitivity to stress, interacting with contextual factors like biological sex, psychological resources, or attachment style.[10–12] The oxytocinergic system is suggested to regulate the stress response in the neuroendocrine and autonomic nervous system, reducing the HPA-axis activity.[13] Based on oxytocin’s buffering effect of stress, a decrease in the expression or sensitivity of the oxytocin receptors – due to *OXTR* SNPs – could allow a greater impact of stress on brain function and structure, and subsequent stress-related psychopathology.

Looking at gene-brain association, we found that the minor allele of the variants *OXTR* rs53576 was associated with reduced right insular brain volumes, which might be a risk factor for FND. While we are not aware of studies that associate the genetic disposition of the oxytocinergic receptor with insular volume, many studies discuss the effect of oxytocin on insular activity. In such, oxytocin related to insular activity seem to be involved in strengthening bottom-up detection of salient signals, which is again sex dependent.[14]

In line with that, we found that for female patients, the *OXTR* rs53576 AA genotype was associated with smaller volumes of bilateral amygdala. Due to the discussed importance of oxytocin and its receptor in the amygdala in regard to fear and anxiety responses,[15] a change in volume of this region might be relevant for the modulation of neural dynamics that are dependent on oxytocin. The sex-specific effect should be investigated, particularly given the predominance of female patients in FND populations,[8] and previous findings suggesting that the *OXTR* rs53576 variant may serve as a protective buffer against stressful experience in a sex-specific way.[10] Stress has been shown to influence amygdalar structure in a manner dependent on timing and sex,[18] which is crucial to consider when discussing the distinct amygdalar alteration observed in FND populations,[19] A more nuanced segmentation in a study with female patients with functional/dissociative seizures further showed a lower right lateral amygdala, along with a higher right central and medial and left anterior amygdala volume. A possible biological explanation for a sex-specific effect of the *OXTR* SNP could be the influence on *OXTR* expression and binding.[20] Overall, these findings underline the importance of considering sex as factors that could significantly interact with the genetic vulnerability for stress-related disorders such as FND.

The major limitation of our study is its small sample size, as genetic studies require large sample size to find relevant clinical-genetic association. Further, using a candidate gene approach and region-of-interest analyses might not allow to identify genetic factors or interaction. However, the candidate gene approach might be appropriate for the given sample size and the exploratory nature of this pilot study, which chose to investigate specific brain regions previously shown to be involved in the pathophysiology of FND. In summary, with oxytocin being one of the regulatory systems upon stress exposure, a decreased sensitivity of the oxytocin receptor due to the *OXTR* genetic variants might exert a greater influence on the stress regulatory system in FND, in a sex-specific manner. Moreover, reduced brain volumes in FND patients might present a biological vulnerability factor with a potential genetic component dependent on sex.

## Data Availability

The data are not publicly available but can be shared upon request.

## ACKNOWLEDGMENTS

We thank all the patients and healthy controls for their participation.

## REFERENCES

1 Perez DL, Nicholson TR, Asadi-Pooya AA, et al. Neuroimaging in Functional Neurological Disorder: State of the Field and Research Agenda. NeuroImage Clin. 2021;30:102623. doi: 10.1016/j.nicl.2021.102623

2 Weber S, Bühler J, Vanini G, et al. Identification of biopsychological trait markers in functional neurological disorders. Brain. 2023;146:2627–41. doi: 10.1093/brain/awac442

3 Spagnolo PA, Norato G, Maurer CW, et al. Effects of TPH2 gene variation and childhood trauma on the clinical and circuit-level phenotype of functional movement disorders. J Neurol Neurosurg Psychiatry. 2020;91:814–21. doi: 10.1136/jnnp-2019-322636

4 Weber S, Rey Álvarez LT, Ansede-Bermejo J, et al. The impact of genetic variations in the serotonergic system on symptom severity and clinical outcome in functional neurological disorders. J Psychosom Res. 2024;186:111909. doi: 10.1016/j.jpsychores.2024.111909

5 Weber S, Heim S, Richiardi J, et al. Multi-centre classification of functional neurological disorders based on resting-state functional connectivity. NeuroImage Clin. 2022;35:103090. doi: 10.1016/j.nicl.2022.103090

6 Weber S, Bühler J, Loukas S, et al. Transient resting-state salience-limbic co-activation patterns in functional neurological disorders. NeuroImage Clin. 2024;41:103583. doi: 10.1016/j.nicl.2024.103583

7 González JR, Armengol L, Solé X, et al. SNPassoc: an R package to perform whole genome association studies. Bioinformatics. 2007;23:654–5. doi: 10.1093/bioinformatics/btm025

8 McLoughlin C, Hoeritzauer I, Cabreira V, et al. Functional neurological disorder is a feminist issue. J Neurol Neurosurg Psychiatry. 2023;94:855–62. doi: 10.1136/jnnp-2022-330192

9 Bartz D, Chitnis T, Kaiser UB, et al. Clinical Advances in Sex- and Gender-Informed Medicine to Improve the Health of All: A Review. JAMA Intern Med. 2020;180:574–83. doi: 10.1001/jamainternmed.2019.7194

10 Byrd AL, Tung I, Manuck SD, et al. An interaction between early threat exposure and the oxytocin receptor in females: Disorder-specific versus general risk for psychopathology and social-emotional mediators. Dev Psychopathol. 2021;33:1248–63. doi: 10.1017/S0954579420000462

11 Notzon S, Domschke K, Holitschke K, et al. Attachment style and oxytocin receptor gene variation interact in influencing social anxiety. World J Biol Psychiatry. 2016;17:76–83. doi: 10.3109/15622975.2015.1091502

12 Saphire-Bernstein S, Way BM, Kim HS, et al. Oxytocin receptor gene (OXTR) is related to psychological resources. Proc Natl Acad Sci U S A. 2011;108:15118–22. doi: 10.1073/pnas.1113137108

13 Takayanagi Y, Onaka T. Roles of Oxytocin in Stress Responses, Allostasis and Resilience. Int J Mol Sci. 2022;23:150. doi: 10.3390/ijms23010150

14 Grace SA, Rossell SL, Heinrichs M, et al. Oxytocin and brain activity in humans: A systematic review and coordinate-based meta-analysis of functional MRI studies. Psychoneuroendocrinology. 2018;96:6–24. doi: 10.1016/j.psyneuen.2018.05.031

15 Huber D, Veinante P, Stoop R. Vasopressin and Oxytocin Excite Distinct Neuronal Populations in the Central Amygdala. Science. 2005;308:245–8. doi: 10.1126/science.1105636

16 Chen FS, Kumsta R, von Dawans B, et al. Common oxytocin receptor gene (OXTR) polymorphism and social support interact to reduce stress in humans. Proc Natl Acad Sci. 2011;108:19937–42. doi: 10.1073/pnas.1113079108

17 Lucas-Thompson RG, Holman EA. Environmental stress, oxytocin receptor gene (OXTR) polymorphism, and mental health following collective stress. Horm Behav. 2013;63:615–24. doi: 10.1016/j.yhbeh.2013.02.015

18 Whittle S, Lichter R, Dennison M, et al. Structural brain development and depression onset during adolescence: a prospective longitudinal study. Am J Psychiatry. 2014;171:564–71. doi: 10.1176/appi.ajp.2013.13070920

19 Hallett M. Functional Neurologic Disorder, La Lésion Dynamique. Neurology. 2024;103:e210051. doi: 10.1212/WNL.0000000000210051

20 Quintana DS, Glaser BD, Kang H, et al. The interplay of oxytocin and sex hormones. Neurosci Biobehav Rev. 2024;163:105765. doi: 10.1016/j.neubiorev.2024.105765

