## Supplemental Material for "Increased Prevalence of Oxytocin Receptor Gene Variant in Functional Neurological Disorder: A Preliminary Case-Control Study"

[e]Centro Nacional de Genotipado (CEGEN), Universidade de Santiago de Compostela, Santiago de Compostela, Spain.

[f]Centre for Biomedical Network Research on Rare Diseases (CIBERER), Instituto de Salud Carlos III, Madrid, Spain.

[g]Instituto de Investigación Sanitaria de Santiago (IDIS), Santiago de Compostela, Spain.

[h]Centro Singular de Investigación en Medicina Molecular y Enfermedades Crónicas (CIMUS), Universidade de Santiago de Compostela, Santiago de Compostela, Spain.

[i]Medicine and Psychiatry Department, University of Cantabria, Santander, Spain.

[j]Fundación Pública Galega de Medicina Xenómica, Sistema Galego de Saúde (SERGAS), Santiago de Compostela, Spain.

#### Questionnaires

All participants completed the Beck Depression Inventory (BDI[1]), the State-Trait Anxiety Inventory (STAI[2]) and the Childhood Trauma Questionnaire (CTQ[3]). For the CTQ, a total CTQ score was calculated by summing the scores of the five subscales.

#### DNA samples

15ml whole-blood was collected from each participant using two 7.5ml EDTA S-Monovette tubes (Sarstedt, Nümbrecht, Germany), and subsequently frozen at -20 °C prior to DNA extraction. DNA was extracted from whole-blood samples using QIAmp DNA Blood kit (Qiagen, Hilden, Germany) according to the manufacturer's protocol. DNA concentration in each sample was then quantified using Quant-it dsDNA Broad-Range Assay Kit (Invitrogen, Thermo Fisher Scientific, Massachusetts, United States) according to the manufacturer's protocol. Each sample was then normalized to 60µl with a concentration of 20ng/µl.

#### Genotyping

10 SNPs were selected based on previous research carried out in other stress-related and mental disorders in association with childhood trauma[4–11]. The selected SNPs are presented in Table 1. Primers were designed using the MassARRAY Assay Design Software and are presented in Supplementary Table 1. DNA samples were genotyped using iPLEX Assay[12] followed by mass spectrometry analysis using the MassARRAY System (Agena Bioscience, San Diego, California). The analysis included the following steps 1) PCR amplification, whereat melting time was adjusted for each primer, 2) Shrimp Alkaline Phosphatase (SAP) treatment to remove unincorporated nucleotides, and 3) single base extension (SBE) to create nucleotide-mass specific products. Processed samples were then dispensed on a 384-element SpectroCHIP bioarray and further analysed using matrix-assisted laser desorption/ionization time-of-flight (MALDI-TOF) technique within the MassARRAY Workstation.

**Supplementary Table 1 Primers used for genotyping**

| Gene Symbol | SNP ID | Reverse primer[a] | Forward primer[a] | SBE primer | Melding temperature (Tm, °C) |
|---|---|---|---|---|---|
| DRD4 | rs3758653 | GAGAAAGTGCTTGCAA AGCG | GAAAATACCTCTCAGG TCAC | GCAAAGCGCAGCAGA GA | 53.8 |
| BDNF | rs6265 | CTTCATTGGGCCGAAC TTTC | GCTTGACATCATTGGC TGAC | CCAACAGCTCTTCTAT CA | 46.2 |
| OXTR | rs2254298 | GCCTGAACAGCTGAAC ATGG | ATTCAGAGGAAGAAGC CCCG | GAAACCATCCCTGTTT TC | 46.1 |
| OXTR | rs53576 | GTAGAATGAGCTTCCC AGCC | TGGAAAGGAAAGGTGT ACGG | TTTCTGTGGGACTGAG GA | 50.5 |
| DRD2 | rs1799732 | AAAGGAGCTGTACCTC CTCG | TCAAAACAAGGGATGG CGGA | CCTCGGCGATCCCCGG CCTG | 64.9 |
| TPH2 | rs4570625 | ACTCACACATTTGCAT GCAC | ACTCATTGACCAACTC CATT | CATTTGCATGCACAAA ATTA | 46.2 |
| FKBP5 | rs1360780 | AGGCACAGAAGGCTTT CACA | TGCCAGCAGTAGCAAG TAAG | GGCTTTCACATAAGCA AAGTTA | 49.8 |
| FKBP5 | rs3800373 | AACCCCTAGTGTAGAA GAGC | TGACTTTTTAGTACTA AGC | AGAGCAACTATTTATT TGTCAAC | 47.6 |
| BDNF | rs1491850 | CCCCATAATTTTACAG CAGG | CCCGAAAGCATATATG CTCC | TGATAATCATACAGAT TTTACGTG | 47.1 |
| TPH1 | rs1800532 | CATGCTCTATATGTGTT AGCC | CAGTGTTACATTCCCT ATGC | ATTATTAATTGACAAC CTATTAGGTG | 47.7 |

[a]The forward and reverse primer sequences contain all the following preceding TAG sequence ACGTTGGATG.

**Supplementary Table 2 Allele frequencies**

| Gene Symbol | SNP ID | Alleles (major/minor) | MAF [%] | *P*-HWE |
|---|---|---|---|---|
| BDNF | rs6265 | C/T | 17.6 | 0.72 |
| | rs1491850 | T/C | 38.2 | 0.82 |
| DRD4 | rs3758653 | T/C | 20.0 | 1.00 |
| DRD2 | rs1799732 | -141 C Ins/Del | 10.0 | 1.00 |
| FKBP5 | rs1360780 | C/T | 31.8 | 1.00 |
| | rs3800373 | A/C | 29.6 | 1.00 |
| OXTR | rs2254298 | G/A | 12.4 | 1.00 |
| | rs53576 | G/A | 42.9 | 0.66 |
| TPH1 | rs1800532 | G/T | 46.5 | 0.51 |
| TPH2 | rs4570625 | G/T | 21.8 | 0.75 |

$P<0.05$ indicates statistical significance. Abbreviations: *BDNF* = brain-derived neurotrophic factor; *DRD4/2* = Dopamine receptor 4/2; *FKBP5* = FK506 binding protein 5; HWE: Hardy-Weinberg equilibrium; MAF: minor allele frequency; *OXTR* = Oxytocin receptor; TPH1/2: Tryptophan hydroxylase 1/2; SNP: Single-nucleotide polymorphism

### Clinical Characteristics patients with FND

**Supplementary Table 3 Clinical Characteristics**

| | FND (N = 85) |
|---|---|
| Disease severity (S-FMDRS, median, quantile) | 6 [2 -13] |
| Duration of illness (in months) | 58.78 (73.0) |
| Symptom type (count)[a] | 44 sensorimotor, 24 gait disorder, 17 tremor, 12 myoclonus, 14 functional/dissociative seizure, 8 dystonia, 7 PPPD, 5 speech disorder, 2 functional deafness, 1 functional vision loss |
| ICD-10 Classification (count)[b] | 62 F44.4, 7 F44.5, 29 F44.6, 8 F44.7, 6 PPPD |
| Psychotropic medication (count) | 14 benzodiazepines, 29 antidepressants, 6 neuroleptics, 9 antiepileptics, 6 opioids |

FND = Functional Neurological Disorder, S-FMDRS = simplified Functional Movement Disorder Rating Scale, PPPD = Persistent Postural-Perceptual Dizziness

[a]Patients can present with several symptom types

[b]Diagnosis of mixed FND (F44.7) was given when F44.4, F44.5, and F44.6 was present

$P^{***} < 0.001$, $P^{**} < 0.01$, $P^{*} < 0.05$.

**Supplementary Table 4 Demographic and clinical data**

| | FND (N = 85) | HC (N = 76) | Statistics |
|---|---|---|---|
| Age, mean (SD), years, [range] | 37.5 (14.2), [17 – 77] | 33.1 (10.9), [18 – 62] | *ns* |
| Sex (females/males) | 63/22 | 55/21 | *ns* |
| Psychotropic medication | 14 benzodiazepines<br>29 antidepressants<br>6 neuroleptics<br>9 antiepileptics<br>6 opioids | 0/76 | *ns* |
| BDI score, mean (SD) | 14.4 (10.0) | 4.59 (6.3) | $Z = -7.55$, $P < 0.0001$ *** |
| STAI-S score, mean (SD) | 37.2 (10.9) | 32.1 (7.7) | $t(132.99) = 7.03$, $P < 0.0001$ *** |
| CTQ total score, mean (SD)[a] | 43.0 (17.0) | 36.3 (13.9) | $Z = -3.03$, $P = 0.002$ ** |
| Emotional Abuse, mean (SD) | 10.1 (5.2) | 8.16 (4.2) | $Z = -2.54$, $P = 0.03$ * |
| Emotional Neglect, mean (SD) | 11.1 (5.2) | 8.80 (4.2) | $Z = -3.09$, $P = 0.01$* |
| Physical Abuse, mean (SD) | 7.15 (3.9) | 5.87 (2.0) | $Z = -2.22$, $P = 0.03$ * |
| Physical Neglect, mean (SD) | 7.68 (3.1) | 6.79 (2.8) | $Z = -2.23$, $P = 0.03$ * |
| Sexual Abuse, mean (SD) | 6.92 (3.9) | 6.72 (3.95) | *ns* |

[a]Childhood trauma questionnaire (CTQ) subscores were corrected for multiple comparison using false-discovery rate (FDR) correction

$P^{***} < 0.001$, $P^{**} < 0.01$, $P^{*} < 0.05$.

### Gene-imaging analysis

**Table 4 Association analysis between *OXTR* rs53576 SNP and brain volumes in FND.**

| Region of interest | Model | Genotype | Full FND sample (N=82) | | | Females only (N=60) | | |
|---|---|---|---|---|---|---|---|---|
| | | | **Cases** | **β* (se)** | ***P*-value** | **Cases** | **β* (se)** | ***P*-value** |
| **Insula right** | Dominant | G/G<br>G/A-A/A | 28<br>54 | 0.00<br>-0.37 (0.1) | 0.04 * | 19<br>41 | 0.00<br>-0.40 (0.14) | 0.1 |
| **Amygdala right** | Recessive | G/G-G/A<br>A/A | 66<br>16 | 0.00<br>-0.04 (0.03) | 0.16 | 47<br>13 | 0.00<br>-0.06 (0.03) | 0.046 * |
| **Amygdala left** | Recessive | G/G-G/A<br>A/A | 66<br>16 | 0.00<br>-0.05 (0.04) | 0.16 | 47<br>13 | 0.00<br>-0.08 (0.04) | 0.02 * |

*A positive beta value refers to higher brain volume for the respective genotype and vice versa. Significance code: *$P$<0.05, **$P$<0.01

#### References


1 Beck AT, Ward CH, Mendelson M, *et al.* An Inventory for Measuring Depression. *Arch Gen Psychiatry*. 1961;4:561–71. doi: 10.1001/archpsyc.1961.01710120031004

2 Spielberger CD, Gorsuch RL, Lushene RE. Manual for the State-Trait Anxiety Inventory. 1970.

3 Bernstein DP, Stein JA, Newcomb MD, *et al.* Development and validation of a brief screening version of the Childhood Trauma Questionnaire. *Child Abuse Negl*. 2003;27:169–90. doi: 10.1016/S0145-2134(02)00541-0

4 Wesarg C, Veer IM, Oei NYL, *et al.* The interaction of child abuse and rs1360780 of the FKBP5 gene is associated with amygdala resting-state functional connectivity in young adults. *Hum Brain Mapp*. 2021;42:3269–81. doi: 10.1002/hbm.25433

5 Womersley JS, Hemmings SMJ, Ziegler C, *et al.* Childhood emotional neglect and oxytocin receptor variants: Association with limbic brain volumes. *World J Biol Psychiatry*. 2020;21:513–28. doi: 10.1080/15622975.2019.1584331

6 Bîlc MI, Vulturar R, Chiş A, *et al.* Childhood trauma and emotion regulation: The moderator role of *BDNF* Val66Met. *Neurosci Lett*. 2018;685:7–11. doi: 10.1016/j.neulet.2018.07.018

7 Ferrer A, Labad J, Salvat-Pujol N, *et al.* BDNF genetic variants and methylation: effects on cognition in major depressive disorder. *Transl Psychiatry*. 2019;9:265. doi: 10.1038/s41398-019-0601-8

8 Spagnolo PA, Norato G, Maurer CW, *et al.* Effects of TPH2 gene variation and childhood trauma on the clinical and circuit-level phenotype of functional movement disorders. *J Neurol Neurosurg Psychiatry*. 2020;91:814–21. doi: 10.1136/jnnp-2019-322636

9 Dadds MR, Schollar-Root O, Lenroot R, *et al.* Epigenetic regulation of the DRD4 gene and dimensions of attention-deficit/hyperactivity disorder in children. *Eur Child Adolesc Psychiatry*. 2016;25:1081–9. doi: 10.1007/s00787-016-0828-3

10 Kohlhoff J, Cibralic S, Hawes DJ, *et al.* Oxytocin receptor gene (OXTR) polymorphisms and social, emotional and behavioral functioning in children and adolescents: A systematic narrative review. *Neurosci Biobehav Rev*. 2022;135:104573. doi: 10.1016/j.neubiorev.2022.104573

11 Funahashi Y, Yoshino Y, Yamazaki K, *et al.* Analysis of methylation and -141C *Ins/Del* polymorphisms of the dopamine receptor D2 gene in patients with schizophrenia. *Psychiatry Res*. 2019;278:135–40. doi: 10.1016/j.psychres.2019.06.001

12 Oeth P, Park C, Kosman D, *et al.* iPLEX$^{TM}$ Assay: Increased Plexing Efficiency and Flexibility for MassARRAY System Through Single Base Primer Extension with Mass-Modified Terminators. 2005.
